# Effectiveness of targeted antenatal family planning information provision on early postpartum family planning uptake in Kisumu county: Study protocol for a Simple Randomized Control Trial

**DOI:** 10.1101/2022.02.18.22271169

**Authors:** Morris Senghor Shisanya, Collins Ouma, Kipmerewo Mary

**Affiliations:** School of Nursing, Kibabii University, Postal Address 1699-50200 Bungoma, Kenya; Department of Biomedical Science and Technology, Maseno University, Private Bag, Maseno, Kenya; School of Nursing, Midwifery and Paramedical Sciences (SONMAPS), Masinde Muliro University of Science and Technology, Postal Address 190-50100, Kakamega, Kenya

**Keywords:** Antenatal family planning information provision, Early Postpartum Family Planning uptake, Fertility intention, Behavioral contracting

## Abstract

Overlooking the contraceptive needs of postpartum women constitutes missed opportunities in health system. Inter-birth interval of at least three years can prevent poor maternal, perinatal and neonatal outcomes and afford women socioeconomic benefits of family planning (FP). The unmet need for FP in the postpartum period remains unacceptably high and far exceeds the FP unmet need of other women. The Kenya Demographic and Health Survey (KDHS) estimate the unmet need for postpartum FP to be 74%. Maternal and Child Health (MCH) continuum provides a great opportunity for postpartum FP (PPFP) interventions integration especially Antenatal targeted FP information giving and gauging fertility intentions. However, there is no protocol for structured, targeted Antenatal FP information giving and behavioural contracting to influence postpartum fertility intentions of mothers before delivery. Knowledge gap regarding fertility intentions and best antenatal strategies for postpartum FP still exists. The available evidence differs across settings and demography. Equally, there has been inadequate exploration of operationally-feasible ways to integrate FP counselling into existing ANC services with limited number of methodologically rigorous trials. This study will therefore examine the effectiveness of antenatal family planning information provision on early postpartum FP uptake using a randomized control trial in Kisumu. The researcher will; assess socio-cultural beliefs towards of PPFP and perceived individual control of PPFP choice, analyze knowledge and intention for PPFP, compare and examine the determinants of PPFP uptake between study groups. A group of 246 antenatal mothers will be randomly assigned to control, community and facility intervention groups as per eligibility criteria in the study facilities that will be cluster sampled. After at least 3 months of intervention and 3 months of postpartum follow-up, clinical superiority will be used to gauge which invention was effective and the model superiority. Questionnaire and Case Report Forms will be the main source of data. The participant will form the unit of analysis which will be by intention to treat. Bivariate analysis will be done as the selection criteria for inclusion of predictors of intention and uptake with a P≤ 0.05 in the final logistic regression model. P-values, ≤0.05, Odds Ratios and 95% confidence interval (CI) will be used to demonstrate significance and the strength of association between selected variables. Dissemination will be through conference presentations and peer reviewed journals. The trial has been registered with the Pan African Clinical Trials Registry PACTR202109586388973 on the 28th September 2021.

## Background

Spacing the inter-birth interval by at least three years by use of effective postpartum contraceptive methods in less developed countries could prevent poor maternal, perinatal, and neonatal health outcomes, including stillbirth, prematurity, low birth weight, neonatal and maternal mortality (1,2).

Ignoring the contraceptive needs of postpartum women constitute missed opportunities in health service delivery to avert unplanned pregnancies and affording every woman and her family the health, social, and economic benefits of family planning (FP) (3).

Globally, approximately 44% of pregnancies are unintended. The unintended pregnancy rates have declined by 30% in developed regions, from 64 per 1000 Women of Reproductive Age (WRA) in 1990s to 45 by 2018. In developing regions, the unintended pregnancy rate reduced by 16% in the same period to 65%. Decline in the unintended pregnancy rate in both regions coincided with increased uptake of family planning (6). Inter-birth intervals in 50% or more of pregnancies in low-income countries are too short at less than 23 months (7). The proportion of postpartum women who no longer want children or want to postpone another child for at least two years but are not using a contraceptive method is still high. The unmet need soon after birth even reaches 75% for the West, and Central Africa region (5).

In Kenya, the situation of PPFP is not different from other low income countries. In a multi-country study which included Kenya, only 25% of women in Kenya had adopted postpartum family planning (PPFP) by six months, and 35% at one year (8). Reproductive, Maternal, Neonatal and Child Health (RMNCH) continuum provides a great opportunity for PPFP interventions (7). Based on the RMNCH program strength in Kenya, Achyut *et al*., (2016) recommended integration of PPFP information giving during ANC period to help in reversing the high unmet PPFP need in Kenya (9). However, Kisumu County, in its strategy for sexual and reproductive health 2019-2024 has overlooked postpartum FP as an intervention of high impact on maternal, new-born and infant outcomes. Such systemic errors point to the extent of the neglect of postpartum family planning locally(10). Equally, the integration of Family Planning information provision in the Antenatal Clinic (ANC) services is not deliberate and the practice is not consistent for every client attending ANC (11)

### Problem Statement

Overlooking the contraceptive needs of postpartum women constitutes missed opportunities in the health care system. Inter-birth interval of at least three years can prevent poor maternal, perinatal and neonatal outcomes and afford women socioeconomic benefits of family planning (FP). The unmet need for FP in the postpartum period remains unacceptably high and far exceeds the FP unmet need of the rest of the women of reproductive age (7,12). Despite observed advances in access to Reproductive, Maternal, Neonatal and Child Health (RMNCH) services in Kenya in recent decades, only 25% of women in Kenya had adopted postpartum family planning (PPFP) by six months, and 35% at one year. This points to feeble progress when it comes to effective postpartum contraceptive services integration (13,14). The study region, Nyanza, posts the shortest median postpartum abstinence with one of the shortest inter-birth intervals nationally. Further, the mean number of children in Kisumu county by the time a woman is 25-40 years is higher (5.6) than the national average (5) (14). Equally, Kisumu County has low (63%) postpartum care provision including PPFP, compared to other Counties that were studied (80%)(11).

### Justification

Routine Antenatal services offer frequent points of contact for providers and pregnant women. These contacts are vital opportunities for information giving, counselling and behavioural contracting to address the contraceptive needs of postpartum women (15). However, there no structured FP information giving, counselling and behavioural contracting to ensure earliest uptake of PPFP and there is still paucity of evidence that this structured integration can work. Likewise, evidence is often weak or incomplete particularly regarding studies that explore the desires, intentions, and priorities of women or couples related to PPFP which may differ across settings and demography. Correspondingly, there has been inadequate exploration of operationally-feasible ways to integrate PPFP information giving and counselling into existing ANC services. This gap of limited number of methodologically rigorous Randomized Control Trials that give a detailed description of tested interventions and of how these were implemented is what this study intends to bridge.

The study will therefore examine the effectiveness of targeted antenatal family planning information provision on Postpartum Family Planning uptake by a rigorous but simple randomized control trial that can be feasibly adopted in ANC practice if proven to be clinically superior. Kisumu County, therefore, will be a suitable site for this study as it has outcomes that point to low early postpartum family planning uptake. Postpartum FP uptake will form a basis for conclusion as to whether the models of PPFP service provision proposed in the trial are clinically superior to the routine practice.

## Methods/Design

### Study Aim

Broadly, the researcher intends to examine the effectiveness of targeted antenatal family planning information provision on early postpartum family planning uptake in Kisumu County. The researcher will specifically: evaluate effect of socio-cultural beliefs on Postpartum Family Planning uptake among postpartum mothers; assess perceived individual control of Postpartum Family Planning choice among postpartum; analyse fertility intentions for postpartum mothers after the intervention; compare Postpartum Family Planning uptake between control and intervention groups of postpartum and examine the determinants of Postpartum Family Planning uptake between control and intervention groups of postpartum mothers in Kisumu County.

### Study design

This will be a prospective interventional study, a Randomised Control Trial (RCT) conducted in Kisumu County, Kisumu East sub County. The randomly sampled facilities are Migosi Health centre and Gita Health Centre for Intervention in urban and rural area respectively. The Community Units (CU) for community-based intervention will be Kuoyo CU and Nyalunya CU in in urban and rural area respectively. The control Facility and CUs will be Kowino HC and Chiga HC and their link CUs in urban and rural area respectively.

The study will have three arms. These will be facility intervention arm, community intervention arm and a control arm. The study will have three interacting phases. The pre-intervention phase, intervention phase and post-intervention phase. The proposed methods in each phase are not complex thus the overall design can be classified as simple intervention design.

Pre-intervention phase is for establishing sampling frame, intervention package and research tools formulation.

The aim of the intervention phase is to determine the effect of the targeted antenatal PPFP information package on the uptake of contraceptive methods during the postpartum period, in comparison with the standard of care. The intervention phase also integrates qualitative research that is aimed at identifying operational barriers and enablers of the intervention outcomes.

Randomization to control and intervention arms is at individual level within randomly selected control, community and the primary level health centres. Participants allocated to the experimental group will receive the targeted antenatal PPFP information package and those allocated to the control group will receive usual antenatal care.

The intervention will be; provision of antenatal information on PPFP using a standardized PPFP counselling tool and postpartum appointment setting. The control group will be under the routine standard antenatal care. There will be antenatal PPFP information provision training for the service providers at the facility and Community Health Assistants (CHA) for the community arm to standardize the intervention.

### Study Population

This study will be among pregnant mothers in their second trimester, irrespective of age, attending ANC clinic in the intervention facilities or are within the respective CUs followed up to 3 months postpartum. The study will be conducted in 2 primary health centres and 1 community in the each of the two sub-counties. The Health centres eligibility depended on if: they offer the continuum of ANC, delivery, and PNC; they provide a selection of at least three modern contraceptive methods that are rated 2 or 1 on the Medical Eligibility Criteria (MEC) for postpartum contraceptives and there were no stock-outs of contraceptives during the preceding six months

### Sampling Method

Multistage sampling will be applied involving purposive sampling for the sub counties, cluster sampling for the intervention and control facilities and community and simple random sampling for the subjects. One sub-county will be purposively sampled by the researcher; one with a rural and an urban set up and should be within easy reach by the team.

Cluster random sampling will then be used to get the 1 facility to be allocated to intervention arm and 1 to the control arm in each of the sub-county. The facilities will be matched based on (1) the average number of deliveries per month and (2) the operational level. Cluster random sampling will also be used to get the intervention CU. Each client meeting the criteria will then be randomly assigned to the study i.e. to the intervention and to the control arms using simple random sampling by picking folded paper labelled “yes” or “no”. There will be no blinding as the intervention facilities and controls are known.

### Sample size determination

The sample size is estimated based on the following assumptions: among women at three months postpartum, KDHS data report 27% of use of any method (modern or traditional) in Kenya while the CPR in the general population is 53% (14). These figures allow the assumption of a desired 26% difference between control and the intervention groups (26% increase in adopting a modern contraceptive by three months postpartum). Therefore the sample size was calculated pairwise for two separate RCT for community arm and the control and the facility arm and the control (16–19). Rosner, 2015 proposed the sample size determination formula for difference in proportions with consideration of type I and II errors and power (18,20), 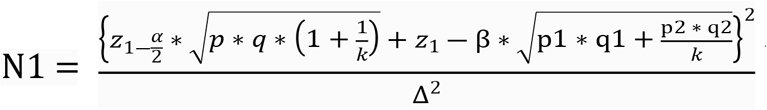 Where *q*_1_ = 1 − *p*_1_, *q*_2_ = 1 − *p*_2_, 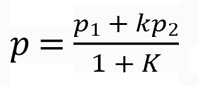, p1, p2 = proportion (incidence) of groups #1 (27%) and #2 (53%), Δ = |p2-p1| = absolute difference between two proportions (0.26), n1 = sample size for group #1, n2 = sample size for group #2, α = probability of type I error (is set at 0.05), β = probability of type II error (is set at 0.1 i.e. 90% power), z = critical Z value for a given α or β(1.96) and K = ratio of sample size for group #2 to group #1(1). Thus for practical equal sample distribution, the actual sample size shall be 246. Each facility shall have 41 clients

### Data collection Procedure

Five tools will be used for data collection, namely; client exit interview guide, case report form, appointment card, Site appraisal form and questionnaire. All the tools will be used to collect quantitative data except site appraisal form and some questions in the questionnaire that need brief explanation. The theory of planned behaviour was applied to design quantitative process and outcome indicators and thus the tools (21). Client exit interview guide and Site appraisal form will be developed based on the procedures set out in the counselling guide. Appointment card will be source of information on client details, proposed date for PPFP initiation and vital PPFP information summary.

### Data collection

Each health centre and community unit will have a trained research assistant. The assistant is to ensure adherence to the study manual and standard operating procedures for data management. The primary outcomes are; PPFP knowledge, the intent for use of PPFP (behavioural contracting) and the actual uptake of modern contraceptive methods at three months postpartum will largely be assessed based on the CRFs, appointment card and questionnaire. Actual uptake of modern contraceptive method will be established three months postpartum between the 12th to 14th weeks after birth. The CRFs will be filled on recruitment by the trained ANC service provider. The appointment card will be filled by the health worker after the client has accepted to set postnatal follow up date for PPFP. The questionnaire will be filled at 14 weeks postpartum during the scheduled MCH visit by a trained enumerator.

A process evaluation will be undertaken with the objectives of understanding the barriers and enablers related to the delivery of PPFP. This will be evaluated based on client exit interviews and site appraisal forms. The client exit interview guide and site visit appraisal form will be used to assess the process quality indicators which will include: waiting time, time/trimester of start of FP counselling, time after FP counselling, group/ individual session, availability of teaching aid on the table during counselling, gauging FP information level, application of GATHER (Greet, Ask, Tell, Help, Explain and Return/Refer) Model and BRAIDED (Benefits, Risks, Alternatives, Inquiries, Decision, Explanation, Documentation) Model. Satisfaction with information given, responses to questions, respectful care, confidentiality and privacy. Client exit interviews will be done by trained enumerator immediately after the intervention has been administered to reduce recall bias by the client. Site appraisal form will be filled on site visits by the research team and on the part of enumerator will be filled each day to include workload for ANC, number of staff available to offer ANC services, availability of FP counselling bag and flipchart.

Internal consistency will be ensured by piloting the tools and refining them to ensure they capture the essence of what they were meant to collect and Cronbach’s alpha of 0.7 will be acceptable.

### Data management

The data will be password protected: only authorized users will be allowed access to the data. In addition, the collected information will be stored safely; hardcopies will be stored in lockable cabinets and soft copies will be secured by password. Data transmission will be encrypted to ensure data integrity and confidentiality of participants.

### Analysis

Quantitative data entry will be done in IBM’s SPSS version 26. The sub-Country research teams will be responsible for verifying the data and a second verification will be done by the researcher to monitor data quality. Questions about data inconsistencies or missing values will be sent to sites and will be resolved on an ongoing basis. The participant will form the unit of analysis and intra-cluster correlation coefficient will be accounted for. All analysis will be by intention to treat Descriptive statistics will be performed by computing means, standard deviations, and minimum and maximum values for continuous variables, and frequencies and percentages for categorical variables. As part of quality control and descriptive analysis of the data, the distribution of variables to detect outliers will be examined. Descriptive statistics will be tabulated for individual clusters and aggregated across clusters.

On evaluating the effect of socio-cultural beliefs on Postpartum Family Planning uptake among postpartum mothers in Kisumu County, descriptive statistics on of prevalent sociocultural beliefs about PPFP will be summarized into frequencies and percentages. Bivariate analysis of effects of sociocultural beliefs on PPFP uptake will be done and presented on two-by-two (2×2) tables with Chi-square being the inferential statistics where P-value will determine the significance of homogeneity of proportions and odds ratio (OR) and 95% confidence interval (95% CI) will demonstrate the strength of the relationship. Binary logistic regression analysis will be done to adjust for confounders of sociocultural beliefs as determinants of PPFP uptake

Perceived individual control of Postpartum Family Planning choice among postpartum mothers in Kisumu County will be analysed using descriptive statistics aggregated based on sociodemographic aspects and presented in tables with means, median, range and standard deviation. This will further be analysed by student t-test to ascertain the significance in differences in means for the determinants of perceived control of family planning choice Significant determinants will be fitted in multilinear regression analysis to adjust for confounders

Fertility intentions for postpartum mothers will be disaggregated based on sociodemographic aspects and other individual characteristics. Intention to use PPFP will be simplified in proportions of the categorical yes or no to appointment for PPFP. Bivariate analysis of determinants of fertility intentions will be done and presented in two-by-two (2×2) tables with Chi-square being the inferential statistics. Binary logistic regression analysis will be done to adjust for confounders of determinants of fertility intentions.

Likewise, level of intention will be measure by Likert scale and analysed by t-test statistics after normality testing and adjusted for in linear regression analysis.

Postpartum Family Planning uptake between control and intervention groups of will be compared by use of simple clinical superiority in the proportions.

Bivariate analysis with chi-square statistics will be used to analyse the determinants of Postpartum Family Planning uptake between control and intervention groups and thus will form the selection criteria for inclusion in final regression model. Binary logistic regression analysis will be applied to adjust for confounders

### Ethical considerations

The study has been approved by Masinde Muliro University of Science and Technology (MMUST) School of Graduate Studies (SGS). Ethical clearance has been obtained from the MMUST Institutional Ethics Review Committee (IERC). An official data collection permission letter will be obtained from the county Director Health and Sanitation. A research authorization and permit has been acquired from NACOSTI. Signed written informed consent for participation will be obtained from all participants after they are introduced to the purpose of the study and informed about their rights. To ensure confidentiality and privacy, the names of the participants will not be recorded in the CRFs and data collection will be in privacy. Principle of justice and impartiality will be adhered to by enabling equal opportunity of the target population to participate in the study by use of probability sampling.

### Trial status

This protocol version number 1 PACTR202109586388973 of the 28th September 2021. The researcher expects recruitment to begin approximately in November 15th 2021 and to be completed by February 2021.

## Data Availability

Type of Access: Controlled Access, Type of analysis: Quantitative analysis Process of Requesting for Data: Request to be done by email of the principal investigator The decision is by the Research team lead by the Principal investigator Criteria for reviewing the request: Qualification of the person requesting, description of the purpose of the request, willingness to engage the Principal investigator in the development of analysis plan, and monographs or manuscripts.

## List of abbreviations

ANC: Antenatal Care
CHA: Community Health Assistants
CPR: Contraceptive Prevalence Rate
CRF: Case Report Form
CU: Community Unit
FP: Family Planning
HC: Health Centre
IBM: International Business Machines Corporation
IERC: Institutional Ethics and Research Committee
PPFP: Postpartum Family Planning
KDHS: Kenya Demographic Health Survey
KNBS: Kenya National Bureau of Statistics
MMUST: Masinde Muliro University of Science and Technology
NACOSTI: National Council of Science and Technology
PNC: Postnatal Care
RCT: Randomized Control Trial
RH: Reproductive Health
RMNCH: Reproductive, Maternal, Neonatal and Child Health
SGS: School of Graduate Studies
SPSS: Statistical Package for Social Sciences
GATHER: Greet, Ask, Tell, Help, Explain and Return/Refer
BRAIDED: Benefits, Risks, Alternatives, Inquiries, Decision, Explanation, Documentation

## Declarations

### Ethics approval and consent to participate

The study has undergone ethics review by Masinde Muliro University of Science and Technology (MMUST), (MMUST/IERC/013/2021) and has been licensed by the National Commission for Science, Technology and Innovation (NACOSTI), Ref. No. 522628. Informed consent will be sought before participant can be recruited.

### Consent for publication

Not applicable

### Availability of data and materials

Reasonable request will be considered by the corresponding author.

Type of Access: Controlled Access,

Type of analysis: Quantitative analysis Process of Requesting for Data: Request to be done by email of the principal investigator The decision is by the Research team lead by the Principal investigator

Criteria for reviewing the request: Qualification of the person requesting, description of the purpose of the request, willingness to engage the principal investigator in the development of analysis plan, and monographs or manuscripts.

### Competing interests

GG is the Chief officer of Health in Kisumu County but is not directly involved in care provision of the essential services to the study participant. The county government of Kisumu nurses employed in the facilities and the Community Health Assistants (CHA) will be the implementers of the intervention.

All other authors declare that they have no conflict of interest.

### Funding

So far, as the protocol manuscript is being prepared, the MSS* will be funding the study for his PhD and is fully responsible for the design, collection, analysis and interpretation of the data and in writing of the manuscript in collaboration with CO and MK.

The County Government of Kisumu will provide space and the nurses and CHAs for implementing the study.

### Authors’ contributions

MSS is the first principal investigator. He conceived the study, led the proposal and protocol development process

CO Contributed to study design and is the lead trial methodologist

MK Contributed to the study design and is lead in public inquiries

## Acknowledgements

Not applicable

## Authors’ information

Morris Senghor Shisanya * Corresponding Author, PhD Student and Lecturer, School of Nursing, Kibabii University, Postal Address 1699-50200 Bungoma, Kenya

Collins Ouma, Professors of Biomedical Sciences, distinguished Member Kenya National Academy of Science (MKNAS),

Department of Biomedical Science and Technology, Maseno University, Private Bag, Maseno, Kenya

Kipmerewo Mary, Professor of Nursing Education, Executive Dean, School of Nursing, Midwifery and Paramedical Sciences (SONMAPS), Masinde Muliro University of Science and Technology, Postal Address 190-50100, Kakamega, Kenya

## SPIRIT Schedule for

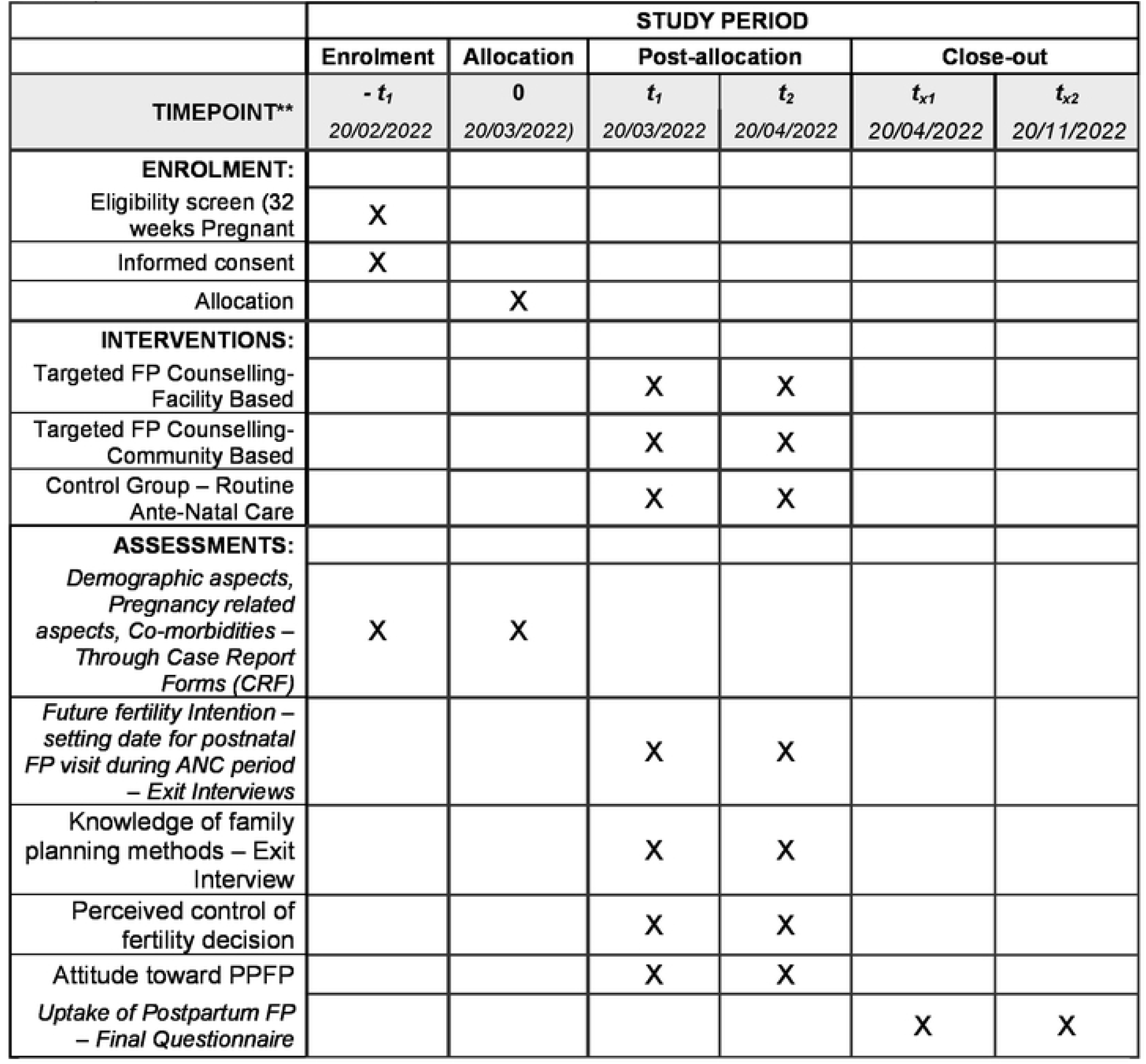

## Notes

### Competing Interest Statement

The authors have declared no competing interest.

### Clinical Trial

PACTR202109586388973 I-decide Project

### Funding Statement

The author(s) received no specific funding for this work.

### Author Declarations

1. Masinde Muliro University of Science and Technology (MMUST) IERC -MMUST/IERC/013/2021 2. National Commission for Science, Technology and Innovation - Ref. No. 522628 3. Consent is written 4. Data will be analyzed anonymously

